# Comparative Study of Point-of-Care Hemoglobin Meters Compared with an Automated Hematology Analyzer in Kumasi, Ghana

**DOI:** 10.64898/2026.03.12.26348261

**Authors:** Amankwaah Lordina, George A. Acheampong

**Author notes:** **Corresponding Author Name:** Lordina Amankwaah, **Address:** Kumasi Technical University, Ghana, **Email:**.

## Abstract

**Background:** Anemia is one of the most prevalent global health challenges, particularly among women of reproductive age and children. Accurate hemoglobin estimation is essential for diagnosing anemia, particularly in resource-limited settings where point-of-care devices are widely used.

**Aim:** To evaluate the diagnostic accuracy of two point-of-care hemoglobin meters compared with a fully automated hematology analyzer.

**Materials and Methods:** A comparative cross-sectional study was conducted among 100 participants at Aniniwaa Medical Centre, Kumasi. Hemoglobin levels were measured using a fully automated analyzer (reference method), Urit and Mission Hb meters. Agreement was assessed using Bland– Altman analysis, and diagnostic performance was evaluated using sensitivity and specificity.

**Results:** The prevalence of anemia was 28% using the analyzer, compared to 60% and 64% using the Urit and Mission meters, respectively. Both meters demonstrated 100% sensitivity but lower specificities (Urit 55.6%; Mission 50.0%). Bland–Altman analysis indicated negative biases (Urit = −1.665 g/dL; Mission = −1.55 g/dL) indicating underestimation of hemoglobin levels.

**Conclusion:** Hemoglobin (Hb) meters offer convenience and portability for field screening but overestimate anemia prevalence. The fully automated analyzer remains more accurate and reliable for diagnosing anemia in clinical settings.

## Introduction

Hemoglobin is a protein in red blood cells that carries oxygen and is composed of a protein called heme, which binds oxygen ^[1, 2]^. Hemoglobin is formed in the red blood cells which is produced in the bone marrow and is made up four polypeptide chains which are two alpha and two beta chains in a normal healthy individual ^[3]^. Hemoglobin estimation indirectly measures the amount of iron in the blood to produce efficient red blood cells ^[4]^. Methods such as the hematocrit method, the drabkin’s solution method, the fully automated method, and point of care testing hemoglobin meters can be used in hemoglobin estimation.

Anemia is a widespread public health issue affecting people across all age groups and is often described as low levels of blood, mainly red blood cells ^[5]^. According to the World Health Organization (WHO), anemia is defined as hemoglobin levels below 13.0 g/dL in adult males and 12.0 g/dL in non-pregnant females and 11.0 g/dL in pregnant women ^[6]^. Since the clinical management of anemia depends on identifying both the presence and severity of hemoglobin reduction, hemoglobin analysis is often the first-line investigation in suspected cases.

In resource-limited settings, hemoglobin meters are mostly used as point-of-care testing for rapid screening of anemia or hemoglobin levels in community and outreach settings because of their portability. However, questions remain about their accuracy and reliability in detecting anemia compared to the fully automated hematology analyzers, which are considered the gold standard in clinical laboratories ^[7]^. According to the National Institutes of Health, hemoglobin (Hb) meters use light absorbance principles to measure Hb concentration in blood, directly or after a chemical reaction that alters the hemoglobin’s light-absorbing properties, and light absorbed is proportional to the hemoglobin concentration ^[4]^.

The fully automated method uses hemoglobin analyzer to estimate hemoglobin levels by using the principle of measuring changes in electrical resistance as cells pass through a sensing area and analysing the scattering of light by cells ^[4]^. Given the widespread use of hemoglobin meters in primary healthcare and outreach programs, it is crucial to assess how their results compare to those from fully automated analyzers.

In remote and low-resource areas where electricity access and laboratory infrastructure is limited, health facilities rely on Hb meters as an alternative to fully automated analyzers ^[8]^. It is essential to assess the agreement between hemoglobin meters and the fully automated analyzer to determine whether Hb meters are reliable methods of Hb estimation. This study will provide valuable information about the performance of Hb meters potentially aiding in better decision making for anemia screening programs in remote and low-resource settings.

## Materials and Methods

### Study Design and Setting

A comparative cross-sectional study was conducted at Aniniwaa Medical Centre, Kumasi, Ghana to evaluate the agreement and diagnostic performance of point-of-care hemoglobin meters against a fully automated hematology analyzer. Aniniwaa Medical Centre is a facility equipped with both point-of-care and automated laboratory systems.

### Study Population

A total of 100 participants referred for routine complete blood count (CBC) testing at Aniniwaa Medical Centre were consecutively recruited.

#### Inclusion Criteria

Consenting patients undergoing CBC testing at Aniniwaa Medical Centre.

#### Exclusion Criteria

Patients with visibly haemolysed or clotted samples.

### Sample Collection and Analysis

Approximately 2 mL of blood was collected into EDTA tube and analyzed within 30 minutes using the fully automated analyzer (Mindray hematology analyzer) and two portable hemoglobin meters (URIT-12 and Mission Hb Meter), using venous aliquots according to manufacturer guidelines. The hemoglobin values from all three methods were recorded in a structured data collection sheet for further analysis.

### Statistical Analysis

Data were analyzed using descriptive statistics. Diagnostic performance was assessed using sensitivity and specificity. Agreement between methods was evaluated using Bland–Altman analysis. A p-value < 0.05 was considered statistically significant.

### Ethical Clearance

The Institute of Research, Innovation and Development in Kumasi Technical University (KsTU) approved the study (Ref No.: IRID/EC2025/HS0041). Verbal informed consent was obtained from all participants and no identifiable features were used in the study.

## Results

### Demographics of Participants

The study recruited a total of 100 participants comprising of 71% females and 29% males. The median age was 26.0 (21.0 – 37.0) years. A summary of the demographic characteristics is shown in **Table 1**.

**Table 1:**
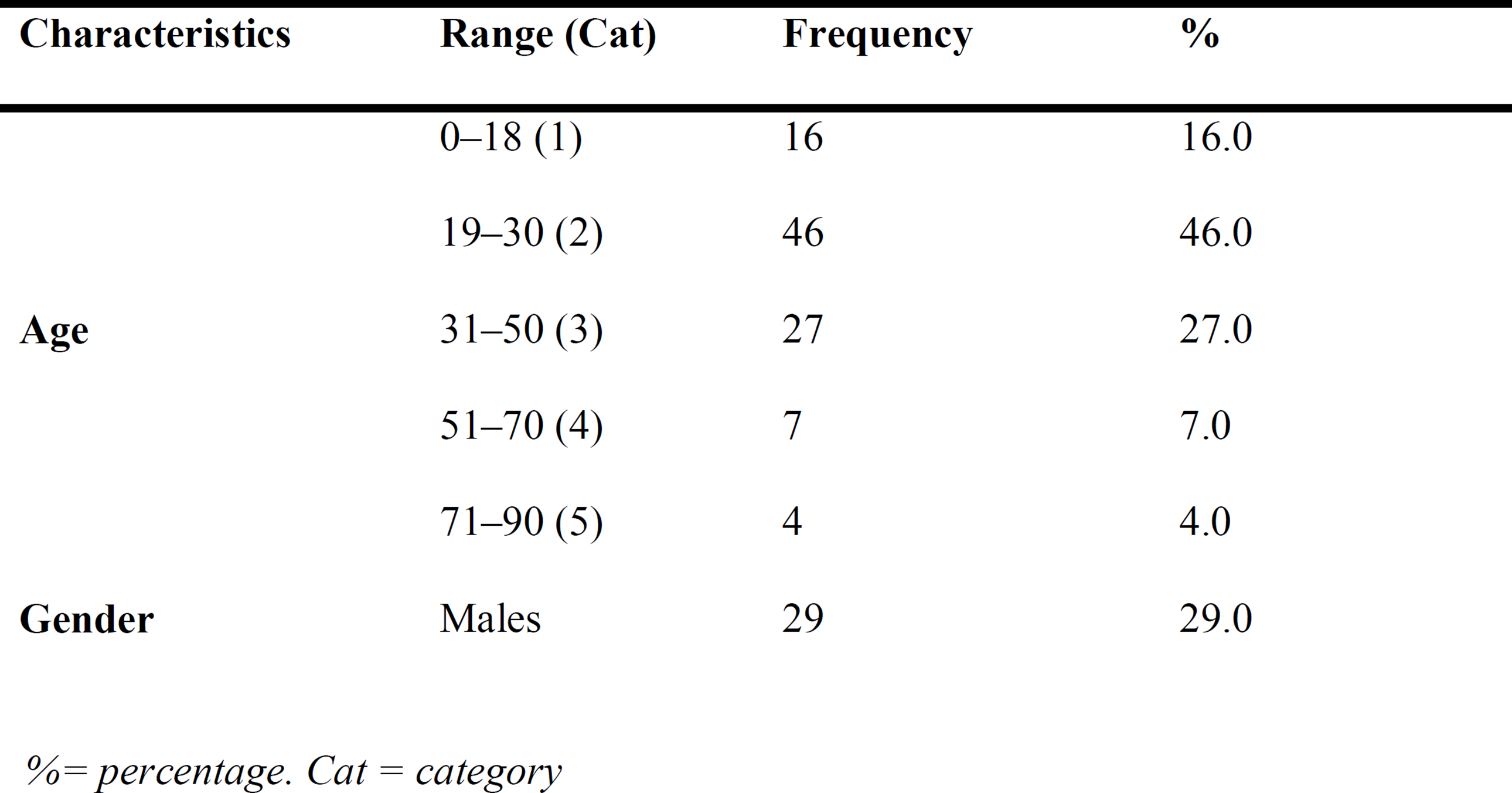
Demographic characteristics of the study participants.

| Characteristics | Range (Cat) | Frequency | % |
| --- | --- | --- | --- |
| Age | 0–18 (1) | 16 | 16.0 |
|  | 19–30 (2) | 46 | 46.0 |
|  | 31–50 (3) | 27 | 27.0 |
|  | 51–70 (4) | 7 | 7.0 |
|  | 71–90 (5) | 4 | 4.0 |
| Gender | Males | 29 | 29.0 |

*%= percentage. Cat = category*

### Prevalence of Anemia as classified by the three estimation methods

Participants with Hb levels less than 11.0 g/dL were classified as anemic. The fully automated analyzer recorded the least prevalence of anemia with a rate of 28% followed by the Urit method classification, 60% and the Mission Hb meter method recorded the highest prevalence of 64% as shown in **Table 2**.

**Table 2:** Prevalence of anemia as classified by the methods of estimation.

| Methods | Anemia |  | Normal |  |
| --- | --- | --- | --- | --- |
|  | Frequency | Percentage (%) | Frequency | Percentage (%) |
| **Urit** | 60 | 60 | 40 | 40 |
| **Mission** | 64 | 64 | 36 | 36 |
| **FAM** | 28 | 28 | 72 | 72 |

*FAM= fully automated analyzer*

### Prevalence of anemia by age and gender using the three estimation methods

The urit meter recorded a high prevalence of anemia in females (73.2%) than males (27.6%). Anemia was prevalent among females in the age range 19–30 (40%) using the Urit meter. **Figure 1** shows the Urit meter prevalence of anemia by age and gender.

**Figure 1:**
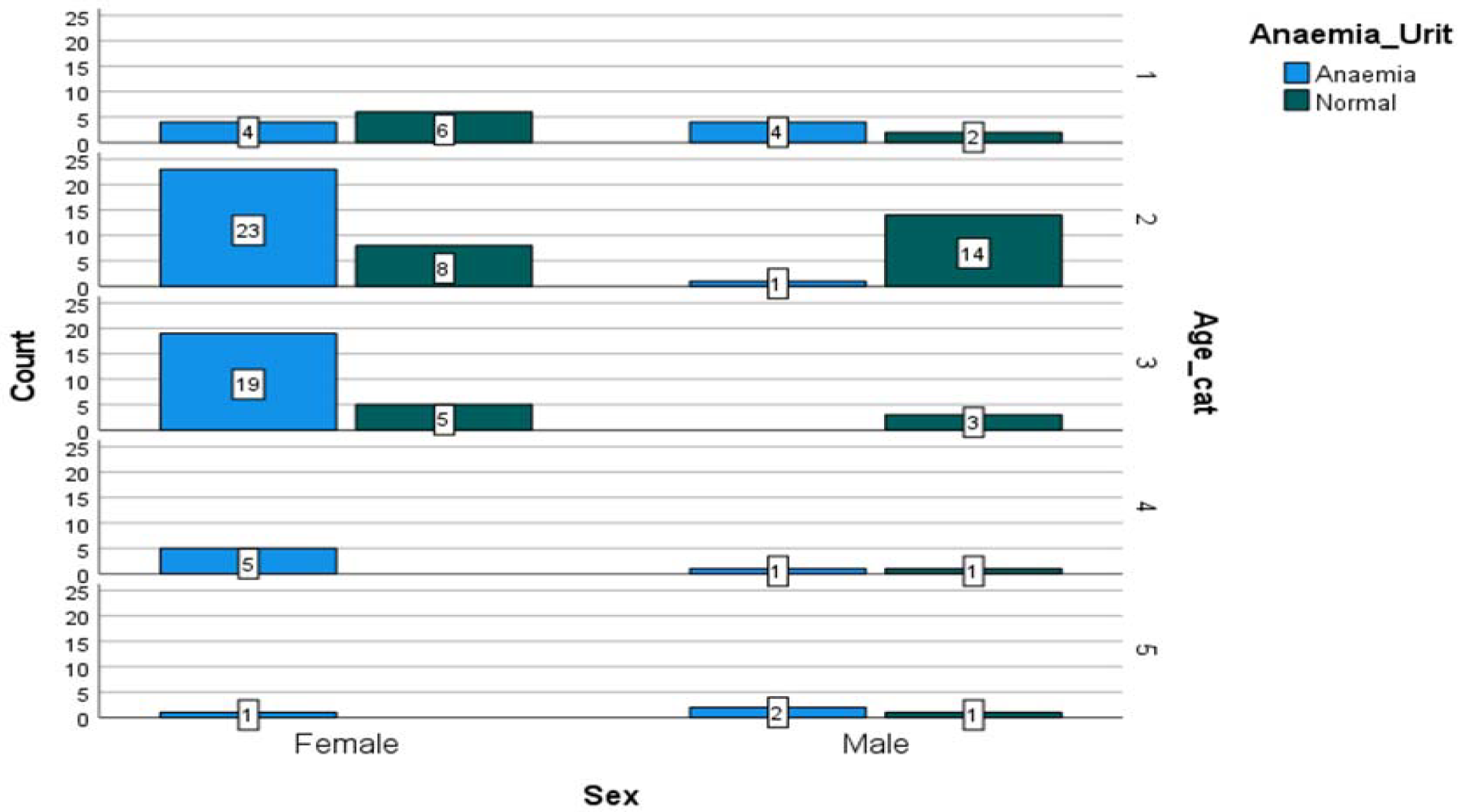
Prevalence of anemia by the Urit Hb meter

The mission hemoglobin meter recorded a high prevalence of anemia in females (76.1%) and anemia incidence was higher in the 31-50 age group (85.2%) as represented in **Figure 2**.

**Figure 2:**
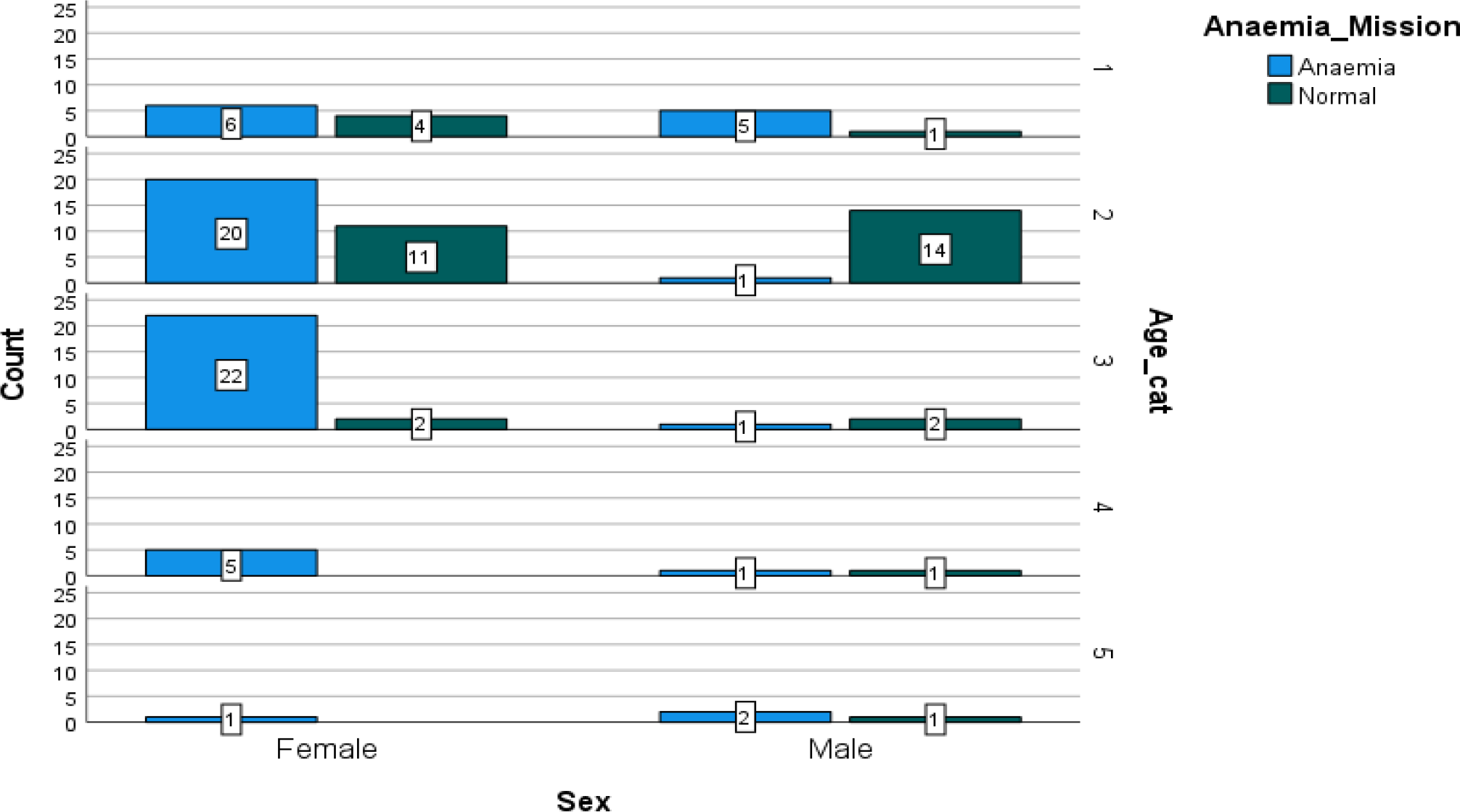
Prevalence of anemia by the Mission Hb meter

The fully automated analyzer (reference) found an anemia prevalence of 36.6% in females and 6.9% in males, shown in **Figure 3**.

**Figure 3:**
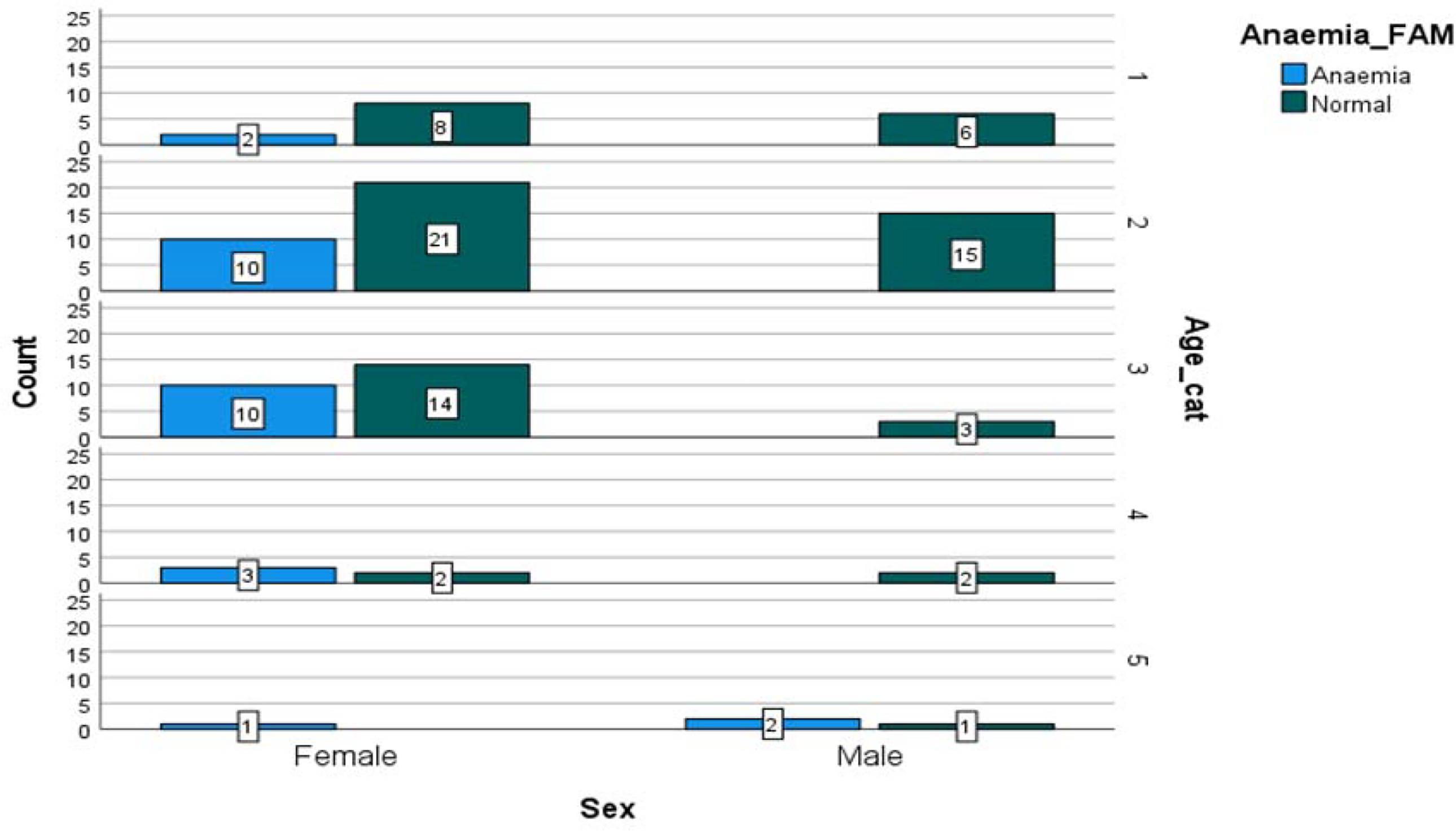
Prevalence of anemia by the fully automated method.

### Assessing the Accuracy of the Hemoglobin meters to the Reference Analyzer Method

**Tables 3 and 4** below demonstrates the accuracy of the Urit and Mission Hb meters to the fully automated analyzer in classifying participants into normal and anemic. The Urit overestimated anemia with a false positive of 32, sensitivity of 100% and a specificity of 55.6%.

**Table 3:**
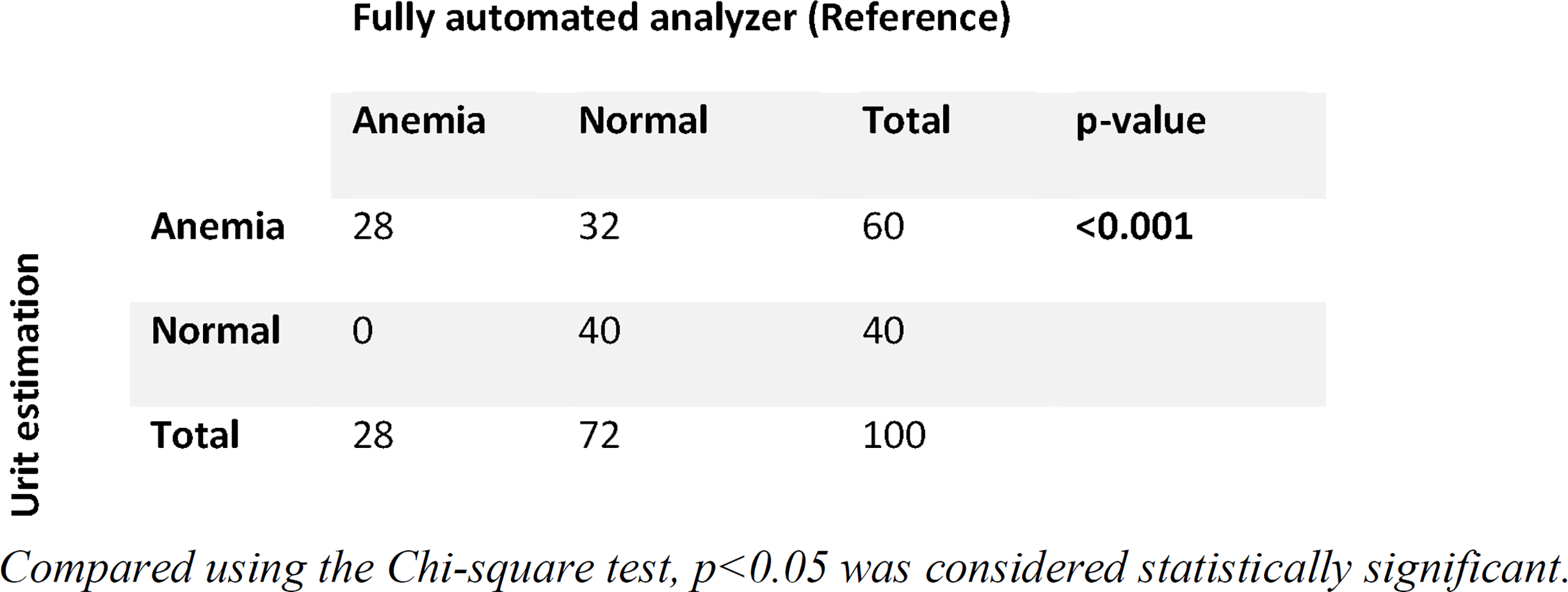
Comparison of anemia status by the Urit and the reference analyzer.

**Table 4:**
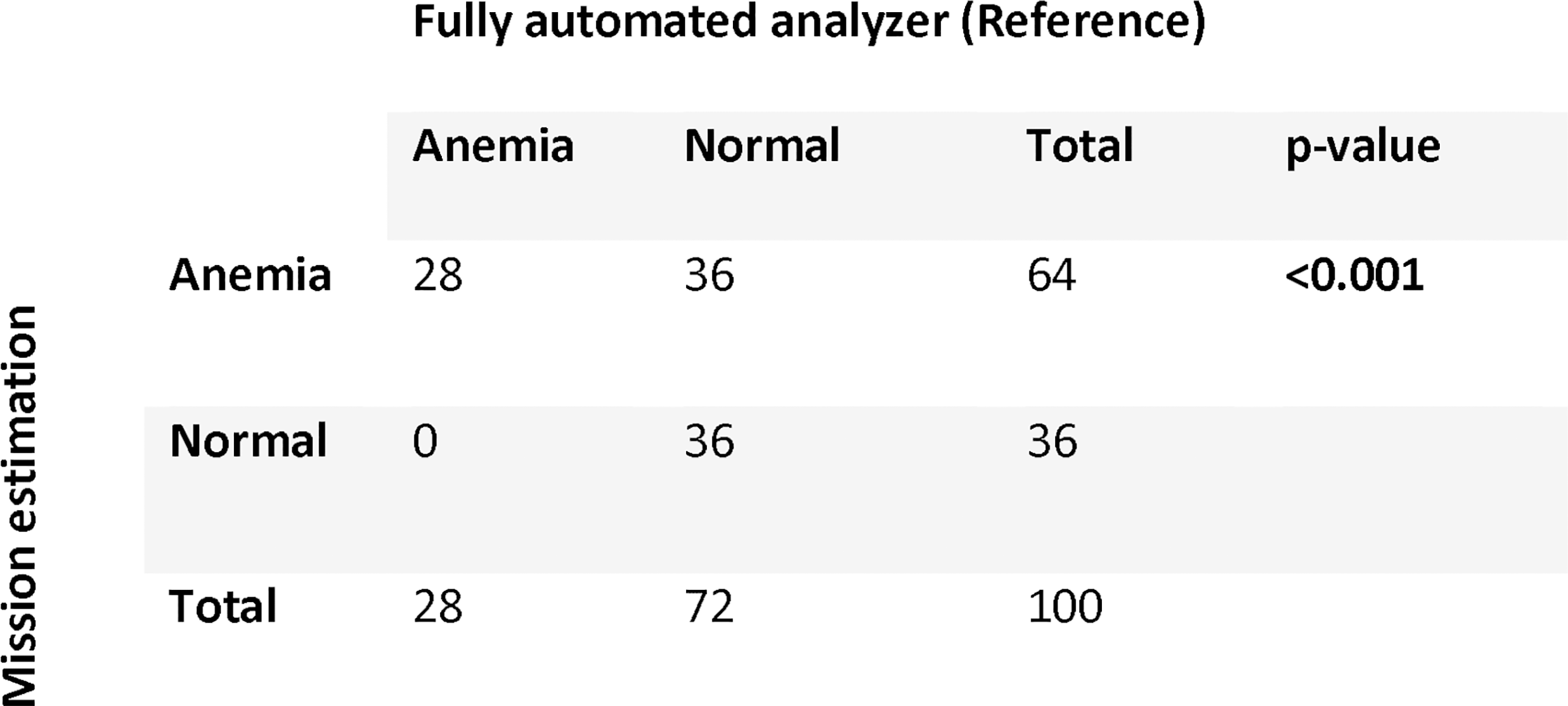
Comparison of anemia status by the Mission and the reference analyzer. Compared using Chi-Square test of independence, p<0.05 considered statistically significant.

The Mission Hb meter recorded a false positive of 36, a 100% sensitivity and showed a specificity of 50% against the standard fully automated analyzer.

### Assessment of the Level of Agreement between the Hemoglobin meters and the Reference Analyzer Method

The Bland-Altman plot was used to determine the level of agreement between the two testing methods (Urit and Mission) and the reference method. The mean bias for Urit meter hemoglobin readings was -01.665 g/dL, with limits of agreement ranging from -4.443 to 1.123 g/dL as shown in **Figure 4**. The Mission hemoglobin meter recorded similar negative bias, with a mean bias of -1.55 g/dL. The lower and the upper limits of agreement were -4.50 and 1.40 g/dL respectively (**Figure 5**).

**Figure 4:**
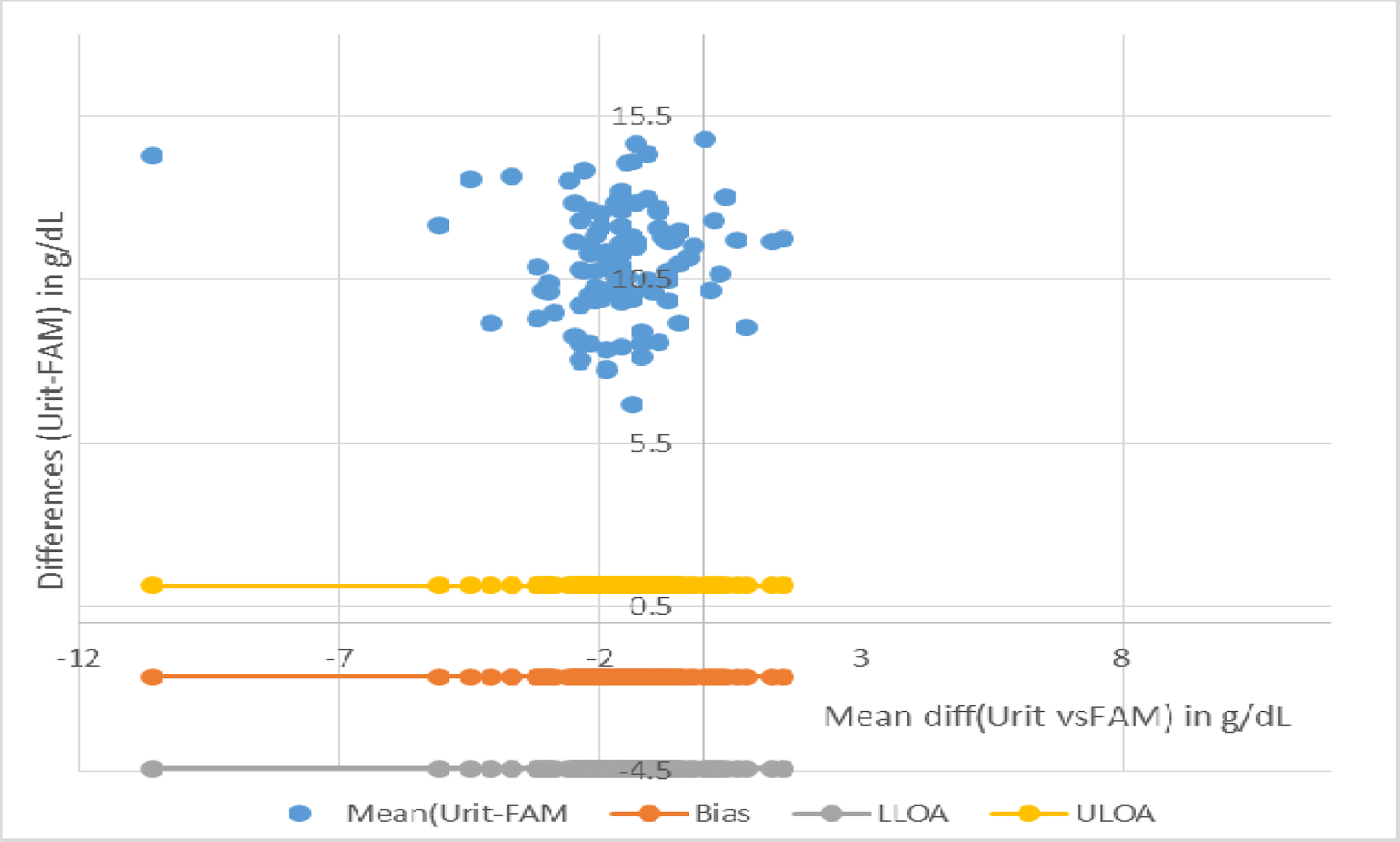
Mean (Urit vs FAM) and Difference (Urit-FAM) Bias= -1.665 Lower limit of agreement (LLOA) = -4.433 Upper limit of agreement (ULOA) = 1.123

**Figure 5:**
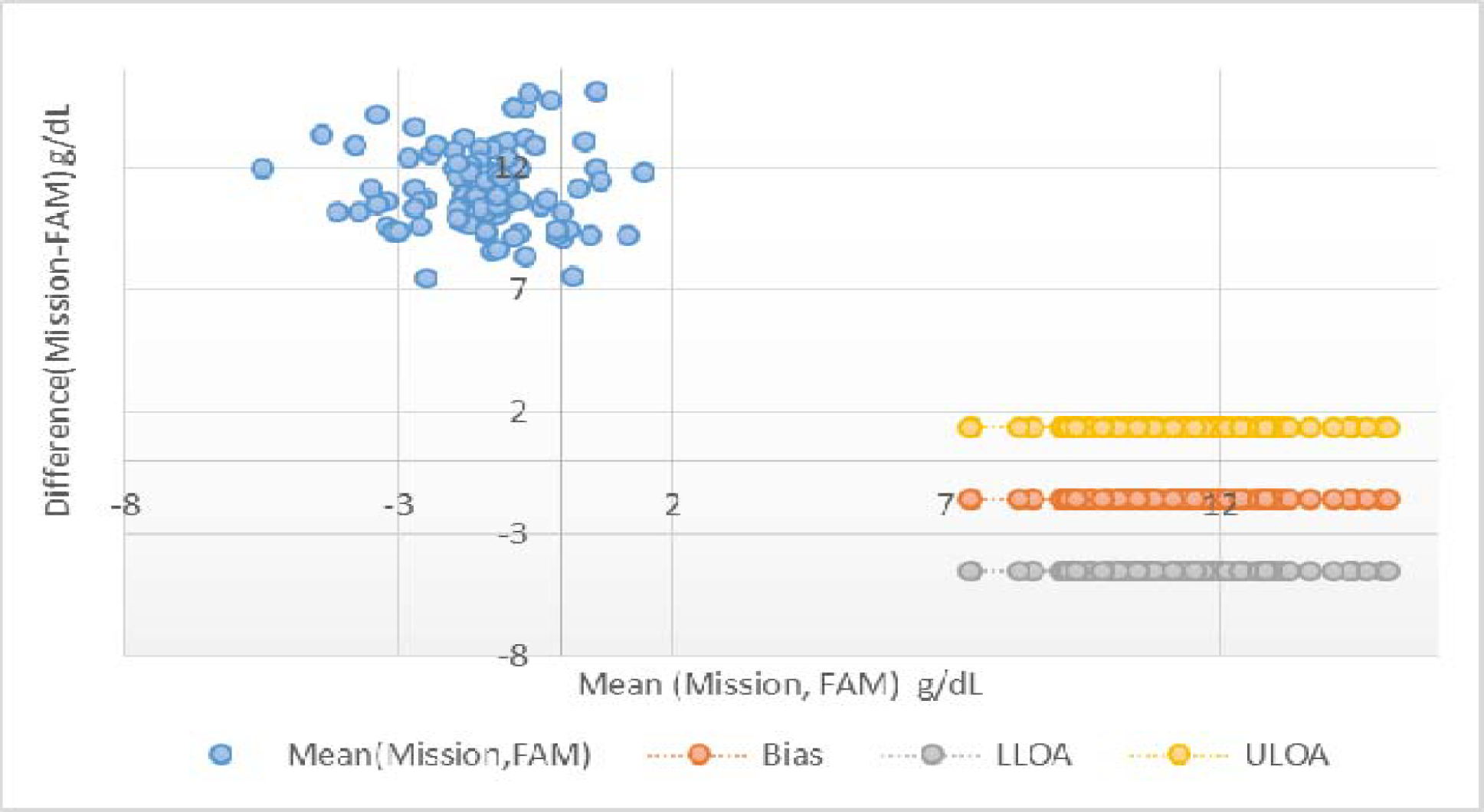
Mean (Mission vs FAM) and Difference (Mission-FAM) Bias= -1.55 LLOA= -4.50 ULOA= 1.40

## Discussion

This study compared the hemoglobin concentrations obtained from the Urit Hb meter, Mission Hb meter, and the Mindray Fully Automated analyzer from 100 participants. The automated analyzer classified fewer participants as anemic compared to the point-of-care (POC) Hb meters. This suggests that the Hb meters may be overestimating the prevalence of anemia, potentially due to device-specific calibration differences or sample type.

Similar findings by ^[8]^ in Ghana observed that POC devices tend to report lower hemoglobin values compared to automated hematology analyzers in malaria-endemic populations, which led to the overestimation of anemia prevalence. In India, ^[9]^ found discrepancies between Hb meters and laboratory analyzers, noting that Bland–Altman plots revealed systematic biases. More recently, ^[10]^ confirmed that while Hb meters remain useful for screening in field surveys, they require strict validation against venous samples analyzed on automated systems to avoid inflated prevalence estimates.

From a public health perspective, these discrepancies are important. If POC devices consistently overestimate anemia prevalence, program planners may allocate resources inefficiently or misinterpret the severity of anemia in a population. However, their portability makes them indispensable tools in rural settings where automated analyzers are unavailable.

### Prevalence of Anemia by gender and age differences

Anemia was classified based on a cut value of <11.0 g/dL. Across methods, females recorded higher anemia prevalence than males with Urit, Mission, and automated analyzer classifying 73.2%, 76.1% and 36.6% of females as anemic respectively. These gender disparities are biologically plausible. In women of reproductive age, iron requirements are elevated due to menstrual blood losses, pregnancies, and lactation ^[11]^. Pregnancy in particular imposes increased demands for iron to support fetal growth and expansion of maternal blood volume, predisposing to anemia when dietary intake or supplementation is inadequate ^[12]^. In Ghana, studies have repeatedly highlighted the burden of anemia in women with ^[13]^ reporting that anemia prevalence among Ghanaian women of reproductive age exceeded 40%, consistent with WHO’s classification of a severe public health problem.

Using the Urit meter, anemia was most prevalent among younger adults (19–30 years), accounting for 40% of all anemic cases. The Pearson Chi-Square test revealed a statistically significant difference with (*X*^*2*^ = 17.881, p<0.001). By contrast, the Mission Hb meter suggested that the highest prevalence occurred among participants aged 31–50 years, with 85.2% classified as anemic in this group (*X*^*2*^= 15.446, p<0.001). The fully automated analyzer reported lower overall prevalence (28%) but similarly identified higher rates among reproductive-aged women (57.1%) (*X*^*2*^= 9.023, p=0.003).

These findings can be understood in relation to life-stage iron requirements and comorbidities. In younger women, menstruation and pregnancy are primary drivers of anemia ^[14]^. This aligns with the Urit meter’s results, where nearly three-quarters of females were classified as anemic. In middle-aged adults (31–50 years), chronic inflammation, liver disease, or parasitic infections (such as hookworm or schistosomiasis) may contribute to anemia in these age groups ^[15]^ as suggested by the Mission Hb meter. Globally, WHO estimates that the burden of anemia peaks in men and women of reproductive age, with prevalence decreasing slightly after menopause in women and remaining consistently low in adult men ^[11]^. The current findings mirror these trends but also suggest device-specific variability in classifying anemia by age group.

### Accuracy of the Hemoglobin meters to the Reference Analyzer Method

The Urit overestimated anemia with a false positive of 32, sensitivity of 100% and a specificity of 55.6%. This difference was statistically significant as shown by the Chi-Square test, (*X*^*2*^= 25.926, *p* <0.05). Similarly, the Mission Hb meter recorded a false positive of 36, a 100% sensitivity and showed a specificity of 50% against the standard fully automated analyzer. The Pearson Chi-Square test showed a statistically significant difference, (*X*^*2*^= 21.875, *p*<0.001).

The assessment of accuracy revealed that both the Urit and Mission Hb meters demonstrated perfect sensitivity (100%) compared with the fully automated analyzer. In practical terms, none of the true anemic cases were missed, which is advantageous in clinical and field screening because it minimizes the risk of failing to detect individuals requiring intervention. However, the Hb meters showed poor specificity suggesting that while the meters are highly reliable in detecting anemia when present, they tend to overestimate its prevalence by misclassifying non-anemic individuals as anemic. Such overestimation was further supported by the statistically significant Chi-square results (p<0.05), confirming that the discrepancy between the Hb meters and the reference analyzer was not due to chance.

This pattern is consistent with earlier studies. ^[9]^ reported that point-of-care Hb meters often over diagnose anemia compared with automated analyzers. In Ghana, ^[8]^ similarly observed that portable Hb meters tended to overestimate anemia prevalence in malaria-endemic areas, cautioning against their use as stand-alone diagnostic tools in epidemiological surveys. Recent evaluations by ^[10]^, also highlighted same limitation, emphasizing the importance of validating point-of-care devices against venous samples using Bland–Altman methods.

The high sensitivity of Hb meters makes them suitable for screening programs where the priority is to detect all potential anemic individuals. However, the low specificity can inflate prevalence estimates and potentially lead to unnecessary supplementation or misallocation of resources. This limitation underscores the importance of using Hb meters as preliminary screening tools rather than definitive diagnostic methods, particularly in research or policy contexts where accurate prevalence data are required.

### Level of Agreement between Urit, Mission, and the Reference Automated Analyzer

The Bland–Altman analysis provided further insight into the level of agreement between the portable Hb meters (Urit and Mission) and the fully automated analyzer. The Hb meters demonstrated a negative mean bias, with the Urit meter showing a bias of –1.665 g/dL and the Mission meter –1.55 g/dL. This consistent underestimation indicates that, on average, the Hb meters reported lower hemoglobin values than the reference analyzer.

The limits of agreement (LOA) for the Urit meter (–4.433 to 1.123 g/dL) and the Mission meter (–4.50 to 1.40 g/dL) were relatively wide indicating that individual readings between the meters and the reference analyzer could differ by as much as ±4 g/dL. Such wide limits raise concerns about reliability at the individual diagnostic level.

These findings align with prior studies that reported systematic underestimation by point-of-care devices. ^[10]^ found that both HemoCue and Mission Hb meters demonstrated mean negative bias when compared with automated hematology analyzers, with similar wide LOAs. In Ghana, ^[8]^ also noted poor agreement between Hb meters and laboratory analyzers, cautioning that while useful for screening, they were less appropriate for precise diagnostics. The negative bias and wide LOA imply that Hb meters overestimate the prevalence of anemia. An individual classified as anemic by a portable meter may not always be confirmed as such by the reference analyzer, highlighting the risk of false positives. Nevertheless, the relatively consistent bias across both Urit and Mission meters shows that systematic calibration adjustments might improve their performance in local contexts.

The Bland–Altman analysis demonstrates that although Hb meters agree reasonably well with the reference method in terms of trend, their underestimation and wide LOA reduce their accuracy for individual-level diagnosis. Confirmatory testing with automated analyzers remains essential, especially in clinical decision-making and epidemiological reporting.

## Conclusion

The Hb meters overestimate anemia prevalence confirmed by their high sensitivity but low specificity, The Bland–Altman analysis further showed that both Hb meters consistently underestimated Hb values compared to the reference analyzer indicating that, while useful for screening, the Hb meters may not provide sufficient accuracy for individual diagnostic purposes without confirmatory testing. While Hb meters provide portable and cost-effective results, their limited accuracy compared to automated analyzers can affect clinical decision-making and public health reporting.

## Data Availability

All relevant data are within the manuscript and its Supporting Information files.

